# Influence of electromagnetic radiation emitted by daily-use electronic devices on the Eyemate^®^ system

**DOI:** 10.1101/19011692

**Authors:** Azzurra Invernizzi, Shereif Haykal, Valeria Lo Faro, Vincenzo Pennisi, Lars Choritz

**Affiliations:** Laboratory for Experimental Ophthalmology, University of Groningen, University Medical Center Groningen, Groningen, the Netherlands; Cognitive Neuroscience Center, Department of Biomedical Sciences of Cells & Systems, University Medical Center Groningen, Groningen, the Netherlands; Department of Ophthalmology, Otto-von-Guericke University Magdeburg, Magdeburg, Germany

**Keywords:** Glaucoma, Intraocular pressure, eyemate^®^ system, telemetry, electromagnetic radiation

## Abstract

**Purpose:** Eyemate^®^ is a system for the continual monitoring of intraocular pressure (IOP), comprised of an intraocular sensor, and a hand-held reader device. The eyemate^®^-IO sensor is surgically implanted in the eye during cataract surgery. Once implanted, the sensor communicates telemetrically with the hand-held device to activate and record IOP measurements. The aim of this study was to assess the possible influence of electromagnetic radiation emitted by daily-use electronic devices on the eyemate^®^ IOP measurements.

**Methods:** The eyemate^®^-IO sensor was placed in a plastic bag, immersed in a sterile sodium chloride solution at 0.9% and placed in a water bath at 37°C. The antenna, connected to a laptop for recording the data, was positioned at a fixed distance of 1 cm from the sensor. Approximately two hours of “quasi-continuous” measurements was recorded for the baseline and for cordless phone, smart-phone and laptop. Repeated measures ANOVA was used to compare any possible differences between the baseline and the tested devices.

**Results:** For baseline measurements, the sensor maintained a steady-state. The same behavior was observed with the devices measurements during active and inactive states.

**Conclusion:** We found no evidence of signal drifts or fluctuations associated with the tested devices, thus showing a lack of electromagnetic interference with data transmission. Patients who already have the eyemate^®^-IO sensor implanted, or those considering it, can be informed that the electromagnetic radiation emitted by their daily-use electronic devices does not interfere with IOP measurements made by the eyemate^®^-IO sensor.

## Background

Glaucoma is one of the leading causes of irreversible blindness worldwide, with a predicted increase in prevalence as the world’s population continues to age.^1^ While the underlying causes of glaucoma vary, the main controllable risk factor for all subtypes of glaucoma is increased intraocular pressure (IOP).^2^ IOP is usually measured by a trained specialist in a clinical setting during working hours. However, IOP exhibits both short and long-term fluctuations throughout the day,^3,4^ which can easily be missed by acquiring static IOP measurements at the clinic in the traditional manner. Although still controversial, some studies have suggested that such fluctuations are an independent risk factor for the development and progression of glaucoma.^5,6^ Therefore, monitoring IOP fluctuations could potentially improve our understanding of glaucoma and how to best control it, and in turn improve patient care.

Several approaches and devices have been proposed for the continuous monitoring of IOP throughout the day.^7^ Currently, the only CE-certified device for continuously monitoring IOP intraocularly is the eyemate^®^ (Implandata Ophthalmic Products GmbH, Hannover, Germany). The eyemate^®^ system comprises a wireless pressure sensor and handheld device (Mesograph). The sensor communicates with the Mesograph device telemetrically, both to provide it with an energy source and to transfer IOP recordings made by the sensor. Implantation of the sensor is usually performed during cataract surgery, where the sensor is placed in the ciliary sulcus after capsular implantation of the intraocular lens. Once implanted, the sensor is meant to remain in the patient’s eye indefinitely.^11^

The eyemate^®^ system enables glaucoma patients to measure their own IOP at any time during the day without the need for a doctor’s visit. It also allows ophthalmologists to produce IOP profiles for their patients throughout the day, enabling the detection of any possible fluctuations. Given the novelty of the device, clinical studies of its long-term outcome are still scarce. A study of the long-term safety of the implanted first-generation sensor in 5 open-angle glaucoma patients over an average period of 37.5 months has reported “good functionality and tolerability”.^8^ A more recent study of patients who received the implant following Boston Keratoprosthesis surgery reported that the sensor successfully detected postoperative IOP peaks and that measurements made by the sensor showed satisfactory agreement with finger palpation.^9^

As the eyemate^®^-IO sensor communicates with the hand-held reader telemetrically, some patients might fear that the electronic devices that they use on a daily basis might somehow interfere with this communication, leading to unreliable measurements of IOP. In this study, we address these concerns by investigating the effect of electromagnetic radiation produced by everyday electronic devices on the measurements made by an eyemate^®^-IO sensor.

## Methods

### Data acquisition

The latest generation of eyemate^®^ wireless intraocular transducer sensor was used for studying the influence of electromagnetic radiation on the IOP measurements. The sensor was placed in a plastic bag, immersed in a sterile sodium chloride solution at 0.9% and placed inside a tissue bath reservoir (RES-01, Experimetria Ltd., Hungary) containing Milli Q-water. A circulating water bath (CWB-02, Experimetria Ltd., Hungary) connected to the tissue bath was used to maintain the temperature constant around 37°C in the system (Figure 1, panel a).

**Figure 1.**
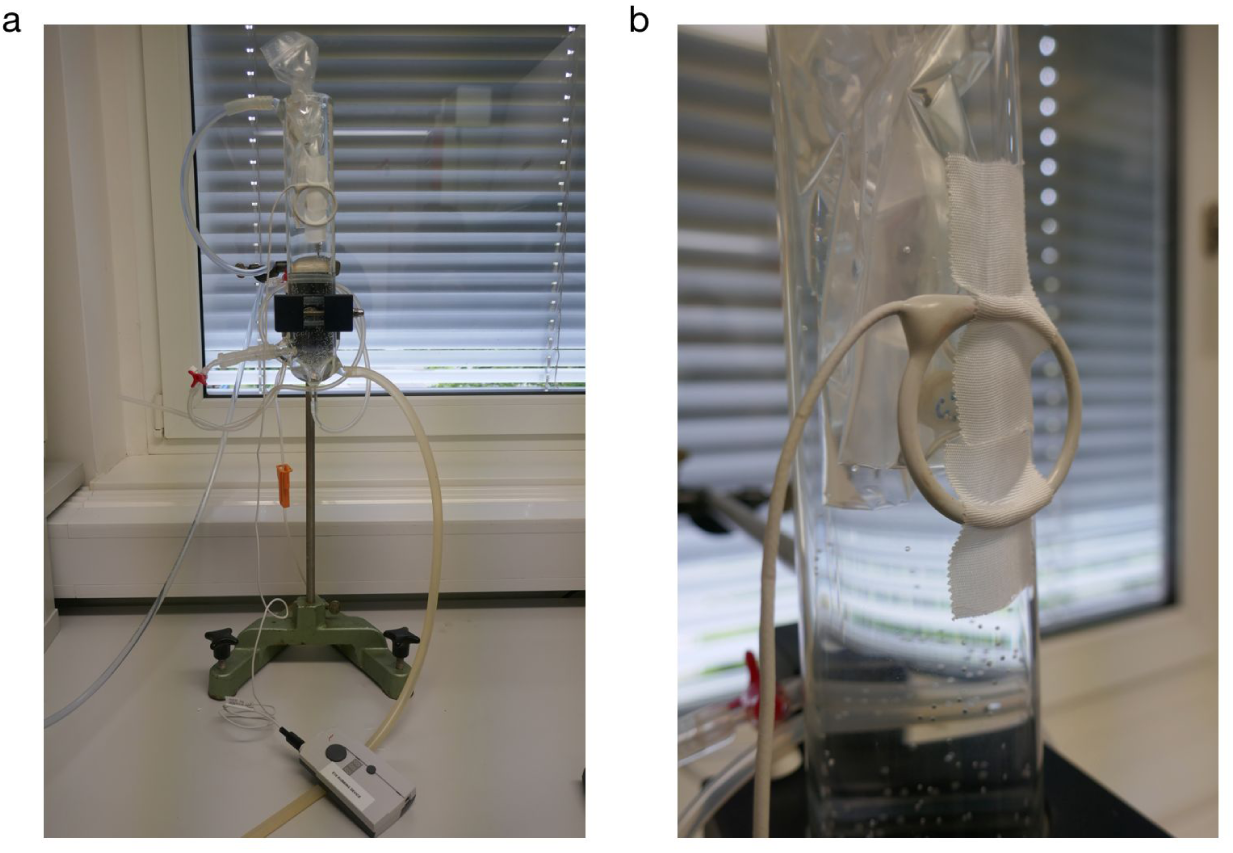
Experimental setup. Panel **a** shows an overview of the setup, including the tissue-bath reservoir, and the Mesograph reader for recording measurements. Panel **b** shows a closer look at the eyemate^®^-IO sensor and the fixed antenna.

An antenna, connected to the Mesograph device, which in turn was connected to a laptop for recording the data, was positioned at a fixed distance of 1 cm from the sensor (Figure 1, panel b). Approximately two hours (116 minutes) of “quasi-continuous” measurements were recorded for the baseline and for each device at a sample rate of approximately 10 Hz. To obtain the baseline measurements, any disturbing electromagnetic impulses were eliminated. All plugs were removed from the sockets in the test room, no lights were switched on, no telephones (fixed line, cordless, or smart-phone). Only the data acquisition computer and the water pump, placed at 2 m from the experimental setup, were left in the room. The duration of the data acquisition was based on the maximal storing capacity of the readout file (64 KB limit).

In order to have comparable measurements for testing the influence of each electronic device, the same environment was recreated: the plugs were removed from the socket, no lights were switched on and only the data acquisition computer and the water pump were left in the room.

For each device, the experiment was divided into four different measurement intervals: the initial twenty-five minutes were used as “baseline” (named no device to avoid confusion); after this time, each single device - smartphone (Huawei P10 Lite), cordless phone (Philips CD180) and laptop (ASUS ZenBook UX410) - was positioned next to the sensor in inactive-mode. For the smartphone and laptop the inactive mode meant to set the device in flight mode while for the cordless phone it meant putting it in stand-by (no call). The measurements in inactive mode were made for twenty-five minutes, after which the device was switched to active mode for the following twenty-five minutes. This consisted of an active call for the smartphone and the cordless phone and active Wi-Fi and video streaming for the laptop. The final measurement period consisted of a no device recording for the remaining forty-one minutes.

### Data analysis

In order to compare the baseline data with data acquired with different devices, the baseline timeline was divided into four sub-phases corresponding to the four measurement intervals (no device, device inactive, device active and no device) acquired for the devices. Absolute pressure values, which represent the raw output data from the sensor used to obtain the final IOP measurement, were used to evaluate possible IOP fluctuation. A polynomial fitting of degree 9th, that takes into account environment pressure and temperature, was applied to the acquired raw data (Figure 2, panel:a). Fitted data was binned by averaging of ten samples per second (Figure 2, panel: b). Using the binned data, we computed the mean, the maximum and minimum values for each measurement period/interval of baseline and devices data. The range of fluctuations for each of the four time-events was then determined by calculating the difference between the maximum and the minimum values per data-bin, then by averaging the max and min values for each event. Repeated measures ANOVA with Greenhouse-Geisser correction using SPSS version 25.0 (SPSS Inc., Chicag, Illinois, USA) was used to statistically compare any possible differences between the baseline and the tested devices to investigate the influence of daily use devices on the sensor recording.

**Figure 2.**
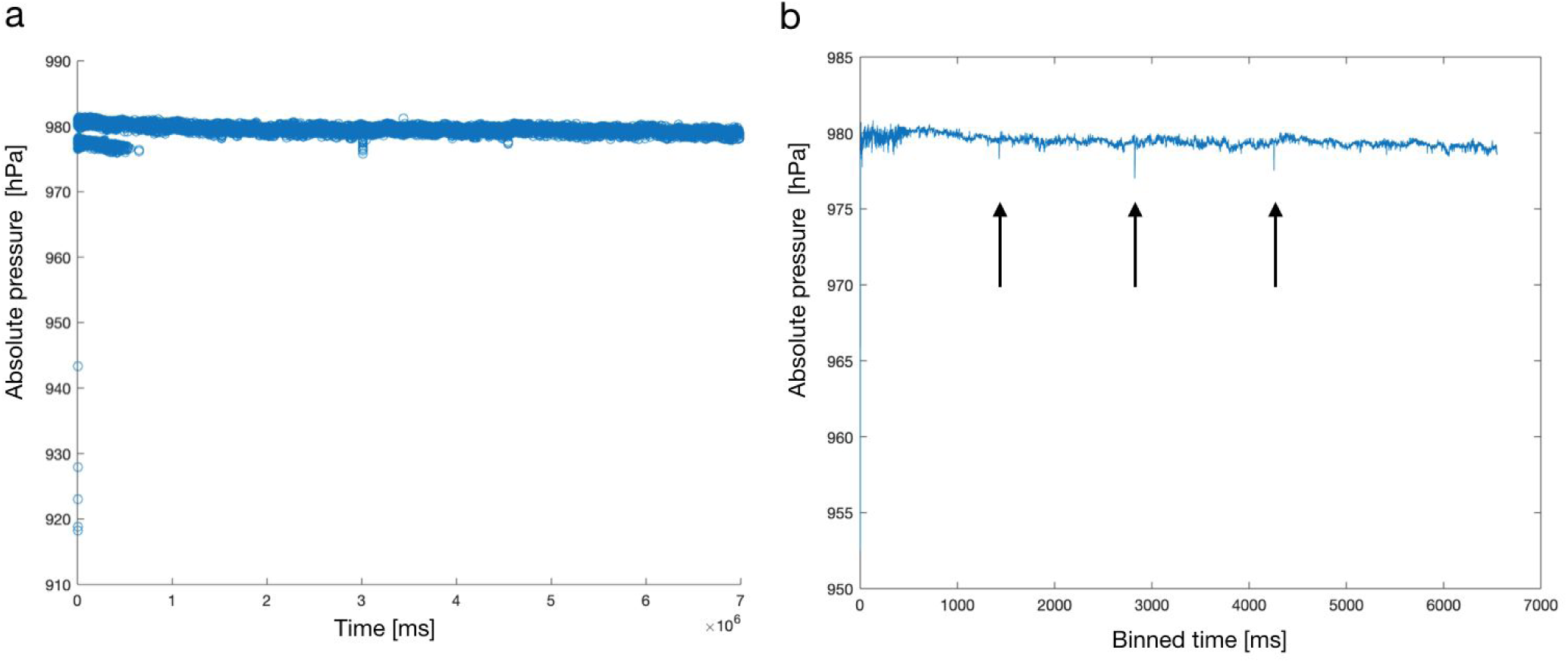
“Quasi-continuous” data recorded for a device. We show an example of the data recorded for one of the three tested devices, namely the smartphone. Panel a: fitted data over time; panel b: fitted and downsampled absolute pressure data over time. Arrows indicate drop in the signal measurements.

## Results

For baseline measurements, the sensor maintained a steady level for the duration of the experiment, resulting in a flat profile with no apparent drift. The same behaviour was observed with the device measurements during active and inactive states.

Small drops in signal measurements were observed corresponding to the time points where each device was handled in the experimental setup (Figure 2, panel: b, drops in different time-events are indicated with arrows).

Similar pattern of distributions and range of fluctuations were observed for both baseline and devices in all four time-events (Figure 3 and Table 1). No statistically significant difference (p-value = 0.332) was found between the average fluctuation for each time-events of the baseline and the tested devices.

**Table 1.** Illustration of absolute pressure range fluctuations during the four time-events. For each time-events, for each device we show the averaged absolute pressure fluctuations calculated based on the range definition.

| Range of fluctuations [hPa] |  |  |  |  |
| --- | --- | --- | --- | --- |
|  | *No Device* | *Device Inactive* | *Device Active* | *No Device* |
| *Baseline* | 0.79 | 0.76 | 0.77 | 0.86 |
| *Phone* | 0.77 | 0.77 | 0.75 | 0.74 |
| *Cordless* | 0.75 | 0.73 | 0.84 | 0.83 |
| *Laptop* | 0.75 | 0.78 | 0.8 | 0.77 |

**Figure 3.**
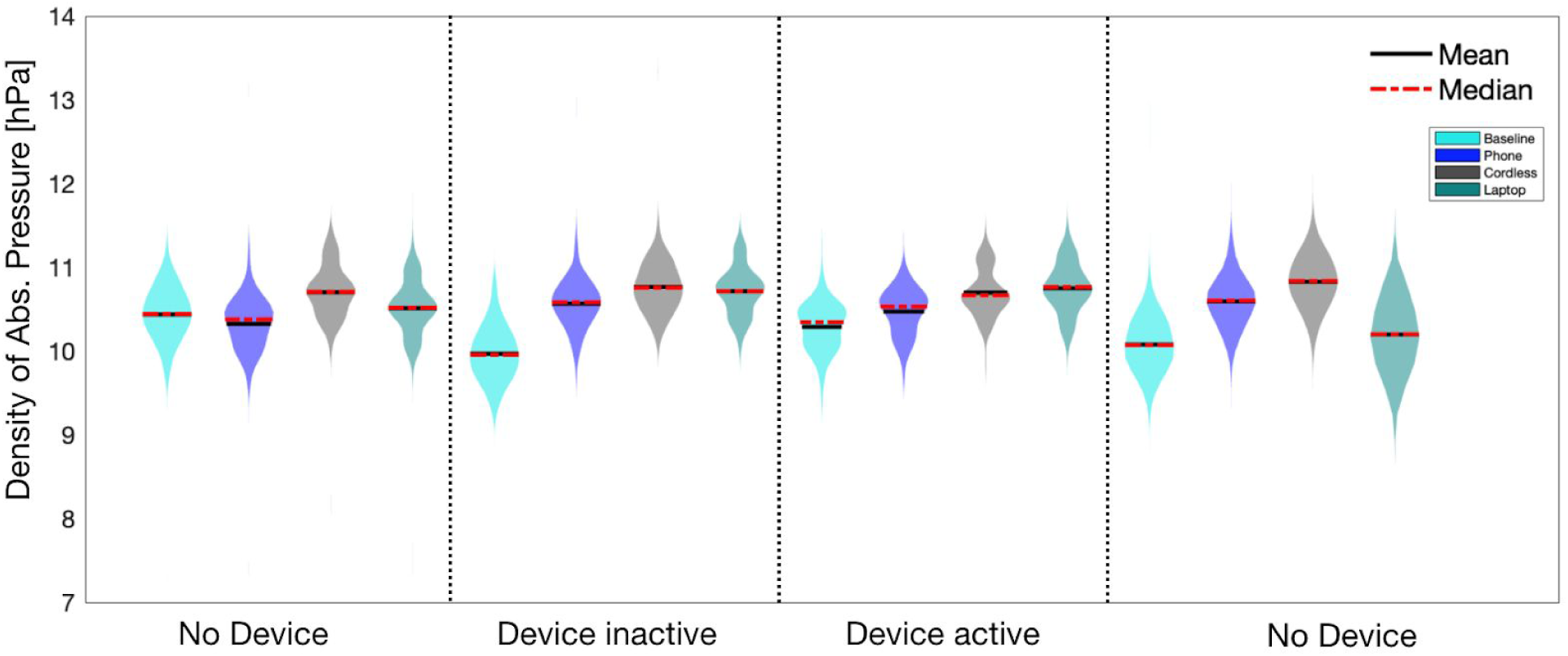
Absolute pressure distributions during the four time-events. A kernel density function was applied to the data for plotting purpose. Mean and median of each distribution are indicated with black solid line and red dotted line, respectively.

## Discussion

The eyemate^®^ is a system capable of continuously monitoring IOP intraocularly. The eyemate^®^-IO sensor is designed to be implanted in the patient’s eye during cataract surgery in order to transmit IOP measurements telemetrically.^10-11^ Because this sensor is constantly sending and receiving electromagnetic signals, patients might fear that their daily use electronic devices may be a source of interference in the IOP measurements. To date, the number of studies investigating this promising new technology and its potential limitations, such as electromagnetic interference, is still lacking.^12-13-14^

Here, we investigated the interference of electromagnetic radiation emitted by three daily-use electronic devices (a cordless phone, a smartphone and a laptop) on the measurements made by the eyemate^®^ system. We found no evidence of signal drifts or fluctuations associated with the tested devices, indicating a lack of interference of the electromagnetic radiation emitted by the devices on the telemetric transmission of data between the sensor and the antenna of the eyemate^®^ system.

However, abrupt signal drops were revealed in the measurement profiles of the three devices, which corresponded with the time points when each device was handled in the experimental setup. These signal drops are most likely unrelated to electromagnetic interference with the readings, but probably due to an abrupt change to the surrounding magnetic field following the repositioning of the tested device close to the sensor. No reduction in the number of samples recorded was present.

A similar study has been previously conducted using the Triggerfish^®^ contact lens sensor (SENSIMED AG, Lausanne, Switzerland) with the same purpose of identifying the influence of electromagnetic radiation on the continuous measurement of the eye pressure by the sensor.^12-13^ The study assessed possible signal drift, noise and fluctuations in IOP measurements recorded by the contact lens sensor due to possible electromagnetic interference from similar daily-use devices. No drift or signal fluctuation was reported.

The Triggerfish^®^ device measures small changes in ocular circumference at the corneal-scleral junction corresponding to changes in intraocular pressure, volume and ocular biomechanical properties as well. Although the Triggerfish^®^ contact lens sensor is placed superficially on the cornea and the eyemate^®^-IO sensor is placed intraocularly, both sensors share a similar method of telemetric communication with an external antenna for IOP monitoring. Therefore, our current results are in line with those reported for the Triggerfish^®^.^13^

### Limitations

An intrinsic limitation of the study is the limited data storage capacity of the reading out system. This restricted our ability to test signal drift and fluctuation over a longer period of time, as was done with similar IOP recording system.^13^ Another possible limitation is the restricted number of devices tested. Future studies may consider including other devices which are becoming more commonly used, e.g. bluetooth headphones, WiFi connected camera.

### Conclusions

Measurements made by the eyemate^®^ system showed no apparent signal drift or evidence of being influenced by external electromagnetic radiation produced by the devices that we tested. Patients who already have the eyemate^®^-IO sensor implanted, and those who are considering to have one, should be informed that the electromagnetic radiation emitted by their daily-use electronic devices does not interfere with IOP measurements made by the eyemate^®^ system.

## Data Availability

data available under request.

## Authors’ contributions

AI, SH, VLF, VP and LC conceptualized, designed the experimental setup and collected the data. AI performed data analysis and visualization. AI, SH and VFL wrote and finalised the manuscript. LC supervised the study and revised the final draft of the manuscript. All authors read and approved the final manuscript.

## Acknowledgements

We want to thank Jacqueline van den Bosch for her useful feedback on the experiment design and Frans W. Cornelissen for his feedback on data analysis and support during the writing process.

